# Virologic and clinical characteristics for prognosis of severe COVID-19: a retrospective observational study in Wuhan, China

**DOI:** 10.1101/2020.04.03.20051763

**Authors:** Sha Fu, Xiao-yu Fu, Yang Song, Min Li, Pin-hua Pan, Tao Tang, Chun-hu Zhang, Tie-jian Jiang, De-ming Tan, Xue-gong Fan, Xin-ping Sha, Jing-dong Ma, Yan Huang, Sha-ling Li, Yi-xiang Zheng, Zhao-xin Qian, Zeng Xiong, Li-zhi Xiao, Hui-bao Long, Jiang-hai Chen, Yi Ouyang

**Author notes:** Address reprint requests to Dr. Yi Ouyang at the Department of Infectious Diseases, Xiangya Hospital, Central South University, Xiangya Rd 87#. Changsha 410008, P.R.China, or at;or to Jianghai Chen, Department of Hand Surgery, Union Hospital, Tongji Medical College, Huazhong University of Science and Technology, Jiefang Avenue 1277#, Wuhan, Hubei 430022, P.R. China, or at. Sha Fu, Xiao-yu Fu, Yang Song contributed equally to this article.

## Abstract

**Background:** The severe acute respiratory syndrome coronavirus 2 (SARS-CoV-2) has progressed to a pandemic associated with substantial morbidity and mortality. The WHO and the United States Center for Disease Control and Prevention (CDC) have issued interim clinical guidance for management of patients with confirmed coronavirus disease (COVID-19), but there is limited data on the virologic and clinical characteristics for prognosis of severe COVID-19.

**Methods:** A total of 50 patients with severe COVID-19 were divided into good and poor recovery groups. The dynamic viral shedding and serological characteristics of SARS-CoV-2 were explored. The risk factors associated with poor recovery and lung lesion resolutions were identified. In addition, the potential relationships among the viral shedding, the pro-inflammatory response, and lung lesion evolutions were characterized.

**Results:** A total of 58% of the patients had poor recovery and were more likely to have a prolonged interval of viral shedding. The longest viral shedding was 57 days after symptom onset. Older age, hyperlipemia, hypoproteinemia, corticosteroid therapy, consolidation on chest computed-tomography (CT), and prolonged SARS-CoV-2 IgM positive were all associated with poor recovery. Additionally, the odds of impaired lung lesion resolutions were higher in patients with hypoproteinemia, hyperlipemia, and elevated levels of IL-4 and ferritin. Finally, viral shedding and proinflammatory responses were closely correlated with lung lesion evolutions on chest CT.

**Conclusions:** Patients with severe COVID-19 have prolonged SARS-CoV-2 infection and delayed intermittent viral shedding. Older age, hyperlipemia, hypoproteinemia, corticosteroid usage, and prolonged SARS-CoV-2 IgM positive might be utilized as predicative factors for the patients with poor recovery.

## Introduction

The novel severe acute respiratory syndrome coronavirus 2 (SARS-CoV-2) that was first reported in Wuhan, China, has caused a large-scale Coronavirus Disease 2019 (COVID-19) pandemic^1^. Currently, more than 823, 626 people have contracted the virus and more than 40, 598 have died worldwide. Robust research is ongoing to help better understand this novel virus^2-7^. However, we still understand little about the virologic characteristics of SARS-CoV-2, and scarce data are available in terms of the risk factors associated with refractory COVID-19.

Currently, interim guidance for discontinuation of transmission-based precautions and disposition of hospitalized patients with COVID-19 have been implemented^8^.

Unfortunately, though, SARS-CoV-2 reactivation and clinical deterioration have been reported in patients after hospital discharge^9^. Besides, some recovered patients can still excrete SARS-CoV-2, as demonstrated by the presence of viral nucleic acids in throat specimens^10^. It can be difficult to detect the virus in recovered patients, and they will be misjudged as “healed” if the patient happens to be at a low level of viral replication. Thus, the potential infectivity of recovered cases remained unclear. A comprehensive exploration of the virologic and clinical characteristics of patients who recovered from COVID-19 would help to address these questions. It also alters us that the current criteria for hospital discharge may need to be re-evaluated^11^.

In this retrospective observational study, 50 severe cases of COVID-19 in Wuhan, China were focused on, and the virologic and clinical characteristics of SARS-CoV-2 were investigated to uncover the natural history of SARS-CoV-2.

## Methods

### Data Source

A single-center, observational study was performed at Union Hospital within the Tongji Medical College of Huazhong University of Science and Technology (HUST). All of the studied adult patients with COVID-19 infection were hospitalized on February 9, 2020. Forty-eight patients were classified as laboratory-confirmed cases of COVID-19 (diagnosis based on positive viral nucleic-acid test results from throat-swab samples), whereas the remaining two patients were clinically diagnosed cases (viral nucleic-acid test was negative, but diagnosis was made based on symptoms, exposure history, and the presence of lung-imaging features consistent with coronavirus pneumonia)^11,12^. Both of the two clinically diagnosed patients were later confirmed using serologic tests for SARS-CoV-2. The medical records from February 9 to March 17, 2020 of the severely ill patients were analyzed. All of the patients were divided into either a good or poor recovery group according to their clinical outcome on March 17, 2020. In addition, the patients were also divided into lung lesions with a partial resolution-patient group and a lung lesions with significant resolution-patient group based on the latest chest CT scans from March 11 to March 17, 2020. This study was approved by the Ethics Commission of Union Hospital at Tongji Medical College of HUST. Written informed consent was obtained directly from patient. The details of the data sources, procedures for nucleic-acids amplification, serologic tests for SARS-CoV-2, treatments, clinical outcomes, and definitions in this study are provided in the supplementary appendix.

### Statistical analysis

For the continuous variables, the descriptive data were expressed as appropriate as the means and standard deviations or the medians and interquartile ranges (IQR). The categorical variables were summarized as the counts and percentages in each category. A Pearson’s χ2 test, a Pearson’s χ2 test with continuity correction, or a Fisher’s exact test were utilized as appropriate to compare the differences between COVID-19 patients with good recovery and poor recovery. All of the analyses were performed using the R statistical package version 3.5.3.

## Results

### General clinical presentations

Fifty patients with COVID-19 were enrolled in this study on February 9, 2020. A total of 27 were male and 23 were female. The median age was 64 years (IQR, 37–87 years). More than half of patients had underlying disorders, including hypertension (10, 20%), diabetes (12, 24%), coronary heart disease (11, 22%), and chronic obstructive pulmonary disease (COPD, [3, 6%]), among other disorders. Most patients experienced symptoms, such as fatigue (39, 78%), fever (28, 56%), and cough (24, 48%). All of the patients had shortness of breath upon admission (Table 1).

**Table 1.** Characteristics of 50 Cases Severely Infected with Covid-19 in Wuhan

|  |  |
| --- | --- |
| **Characteristic** | **Value** |
| **Age** |  |
| Median (range) — yr | 64 (37-87) |
| **Subgroup — no. (%)** |  |
| 37–49 yr | 6 (12.0) |
| 50–65 yr | 23 (46.0) |
| 66–75 yr | 15 (30.0) |
| 76–87 yr | 6 (12.0) |
| **Male sex — no. (%)** | 27 (54.0) |
| **Familial cluster — no. (%)** |  |
| 1 | 25 (50.0) |
| 2 | 13 (26.0) |
| 3 | 9 (18.0) |
| 4 | 1 (2.0) |
| 6 | 2 (4.0) |
| **Smoking history — no. (%)** | 10 (20.0) |
| **Comorbidity— no. (%)** |  |
| Diabetes | 12 (24.0) |
| Coronary heart disease | 11 (22.0) |
| Hypertension | 10 (20.0) |
| Chronic obstructive pulmonary disease | 3 (6.0) |
| Chronic liver disease | 2 (4.0) |
| Chronic kidney disease | 1 (2.0) |
| Others | 6 (12.0) |
| **Fever — no. (%)** |  |
| 37.3–38.0°C | 9 (18.0) |
| 38.1–39.0°C | 11 (22.0) |
| >39.0°C | 8 (16.0) |
| **Symptoms — no. (%)** |  |
| Shortness of breath | 50 (100.0) |
| Fatigue | 39 (78.0) |
| Loss of appetite | 33 (66.0) |
| Dizzy | 29 (58.0) |
| Cough | 24 (48.0) |
| Somnopathy | 16 (32.0) |
| Diarrhea | 11 (22.0) |
| Sputum production | 10 (20.0) |
| Nausea or vomiting | 7 (14.0) |
| Myalgia or arthralgia | 6 (12.0) |
| Conjunctivitis | 1 (2.0) |
| Sore throat | 1 (2.0) |
| **Coexisting disorder — no. (%)** |  |
| Electrolyte disturbance | 43 (86.0) |
| Coagulopathy | 39 (78.0) |
| Hyperlipidemia | 33 (66.0) |
| Hypoproteinemia | 30 (60.0) |
| Acute liver injury | 21 (42.0) |
| Acute kidney injury | 4 (8.0) |
| Acute cardiac injury | 8 (16.0) |
| Secondary infection | 1 (2.0) |
| **Laboratory findings** |  |
| Leukocyte count — no. (%) |  |
| Mean count — per mm<sup>3</sup> | 5496±1833 * |
| <4,000/mm<sup>3</sup> | 9 (18.0) |
| >10,000/mm<sup>3</sup> | 2 (4.0) |
| Lymphocyte count — no. (%) |  |
| Mean count — per mm<sup>3</sup> | 1208±475 * |
| <1,500/mm<sup>3</sup> | 41 (82.0) |
| Neutrophil: lymphocyte ratio >2.5 | 34 (68.0) |
| Platelet count — no. (%) | 222.34±112.80 * |
| <150,000/mm<sup>3</sup> | 16 (32.0) |
| >350,000/mm<sup>3</sup> | 9 (18.0) |
| CD4 T-lymphocyte count — per mm<sup>3</sup> |  |
| Mean count — per mm<sup>3</sup> | 43.98±8.63 * |
| CD8 T-lymphocyte count — per mm<sup>3</sup> |  |
| Mean count — per mm<sup>3</sup> | 25.98±7.47 * |
| CD4:CD8 ratio <1.4 — no. (%) | 16 (32.0) |
| **Inflammatory biomarkers — no. (%)** |  |
| Erythrocyte sedimentation rate >20 mm/hour | 43 (86.0) |
| Ferritin >274.66 ng/ml | 32 (64.0) |
| C-reactive protein >8 mg/liter | 21 (42.0) |
| High-sensitivity C-reactive protein >8 mg/liter | 12 (24.0) |
| Procalcitonin ≥0.5 ng/ml | 1 (2.0) |
| Interleukin-6 >2.9 pg/ml | 45 (90.0) |
| Interleukin-2 >3.2 pg/ml | 20 (40.0) |
| Interleukin-4 >3.2 pg/ml | 20 (40.0) |
| Interleukin-10 >5.0 pg/ml | 8 (16.0) |
| **Blood biochemical — no. (%)** |  |
| Cholesterol >5.2 mmol/liter | 31 (62.0) |
| Triacylglycerol >1.7 mmol/liter | 23 (46.0) |
| Albumin <35 g/liter | 30 (60.0) |
| Lactate dehydrogenase >250 U/liter | 11 (22.0) |
| Alanine aminotransferase >40 U/liter | 12 (24.0) |
| Aspartate aminotransferase >40 U/liter | 10 (20.0) |
| High-sensitive cardiac troponin >26.2 ng/liter | 8 (16.0) |
| **Abnormal coagulation findings — no. (%)** |  |
| D-dimer >0.5 mg/liter | 31 (62.0) |
| Fibrinogen >4 g/liter | 22 (44.0) |
| **Minerals — no. (%)** |  |
| Anion gap <8 mmol/liter | 42 (84.0) |
| Potassium <3.5 mmol/liter | 12 (24.0) |
| **Abnormalities on chest CT — no. (%)** |  |
| Ground-glass opacity | 49 (98.0) |
| Bilateral patchy shadowing | 49 (98.0) |
| Multifocal | 46 (92.0) |
| Centrilobular nodules | 37 (74.0) |
| Peripheral | 22 (44.0) |
| Consolidation | 21 (42.0) |
| Pleural thickening | 21 (42.0) |
| Mediastinal lymphadenopathy | 21 (42.0) |
| Subpelural line | 12 (24.0) |
| Fibrosis | 9 (18.0) |
| **Treatment — no. (%)** |  |
| Antibiotics | 33 (66.0) |
| Antiviral drugs | 50 (100.0) |
| Human serum albumin | 23 (46.0) |
| Corticosteroid | 14 (28.0) |
| Gamma globulin | 9 (18.0) |
| Tocilizumab | 6 (12.0) |
| Thymosin | 50 (100.0) |
| **Oxygen therapy method — no. (%)** |  |
| Nasal catheter | 38 (76.0) |
| Mask | 12 (24.0) |
| **FiO<sub>2</sub>(L/min) — no. (%)** |  |
| <5 | 27 (54.0) |
| ≥5 | 23 (46.0) |

\* These patients were all residents of Wuhan. The median age of the patients was 64 years (range, 37 to 87 years). Percentages may not total 100 because of rounding. Familial cluster represents the number in a family who were infected COVID-19 at the same time.
\* Plus-minus values are means ± SD. The definitions of coexisting disorder in this study was provided in the Supplementary Appendix. CT denotes computed tomography.

Most patients exhibited lymphopenia upon admission. The common complications while in the hospital were electrolyte disorders, coagulopathy, dyslipidemia, hypoproteinemia, and liver dysfunction. Importantly, hyperlipidemia, hypoproteinemia, and liver injury were more frequently observed in patients with poor recovery, with TG and lactatdehydrogenase (LDH) levels above the normal range. In addition, coagulation disorders were commonly seen in patients with poor recovery, and the frequency of fibrogen (FIB) elevation differed greatly between the two groups (Table 2).

**Table 2.**
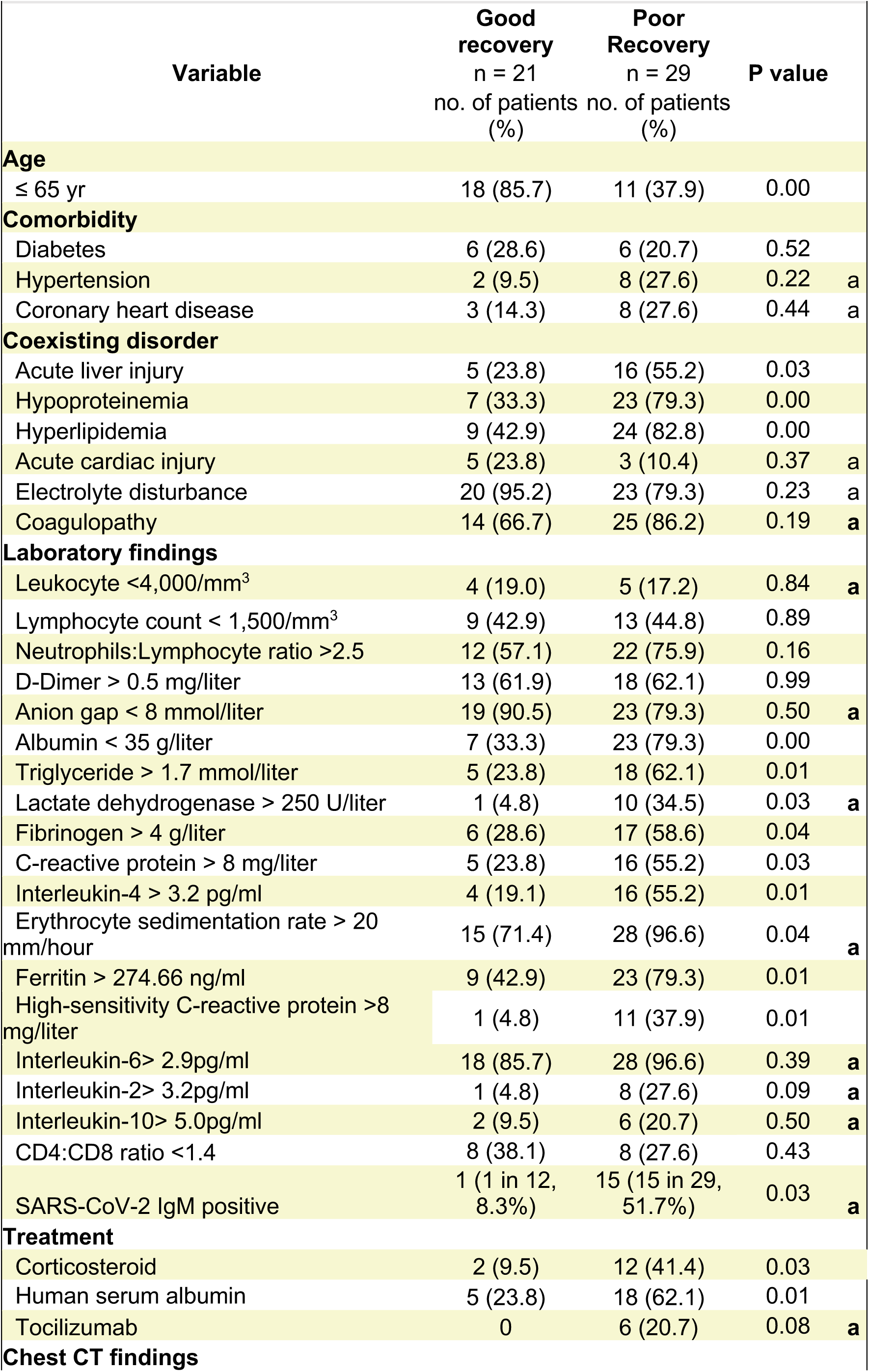
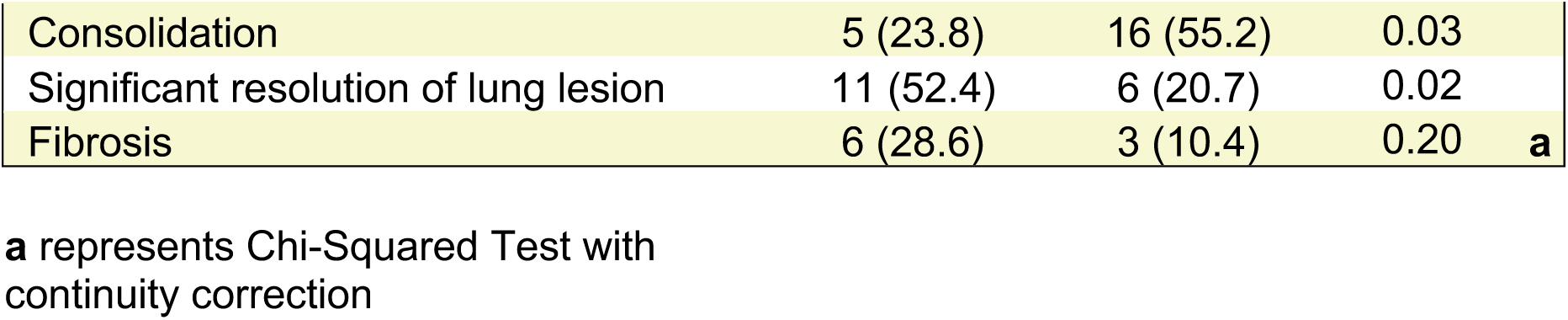
Clinical Characteristics of Good Recovery Patients and Poor Recovery Patients

| Variable | Good recovery<br>n = 21<br>no. of patients (%) | Poor Recovery<br>n = 29<br>no. of patients (%) | P value |
| --- | --- | --- | --- |
| **Age** |  |  |  |
| ≤ 65 yr | 18 (85.7) | 11 (37.9) | 0.00 |
| **Comorbidity** |  |  |  |
| Diabetes | 6 (28.6) | 6 (20.7) | 0.52 |
| Hypertension | 2 (9.5) | 8 (27.6) | 0.22 a |
| Coronary heart disease | 3 (14.3) | 8 (27.6) | 0.44 a |
| **Coexisting disorder** |  |  |  |
| Acute liver injury | 5 (23.8) | 16 (55.2) | 0.03 |
| Hypoproteinemia | 7 (33.3) | 23 (79.3) | 0.00 |
| Hyperlipidemia | 9 (42.9) | 24 (82.8) | 0.00 |
| Acute cardiac injury | 5 (23.8) | 3 (10.4) | 0.37 a |
| Electrolyte disturbance | 20 (95.2) | 23 (79.3) | 0.23 a |
| Coagulopathy | 14 (66.7) | 25 (86.2) | 0.19 a |
| **Laboratory findings** |  |  |  |
| Leukocyte <4,000/mm<sup>3</sup> | 4 (19.0) | 5 (17.2) | 0.84 a |
| Lymphocyte count < 1,500/mm<sup>3</sup> | 9 (42.9) | 13 (44.8) | 0.89 |
| Neutrophils:Lymphocyte ratio >2.5 | 12 (57.1) | 22 (75.9) | 0.16 |
| D-Dimer > 0.5 mg/liter | 13 (61.9) | 18 (62.1) | 0.99 |
| Anion gap < 8 mmol/liter | 19 (90.5) | 23 (79.3) | 0.50 a |
| Albumin < 35 g/liter | 7 (33.3) | 23 (79.3) | 0.00 |
| Triglyceride > 1.7 mmol/liter | 5 (23.8) | 18 (62.1) | 0.01 |
| Lactate dehydrogenase > 250 U/liter | 1 (4.8) | 10 (34.5) | 0.03 a |
| Fibrinogen > 4 g/liter | 6 (28.6) | 17 (58.6) | 0.04 |
| C-reactive protein > 8 mg/liter | 5 (23.8) | 16 (55.2) | 0.03 |
| Interleukin-4 > 3.2 pg/ml | 4 (19.1) | 16 (55.2) | 0.01 |
| Erythrocyte sedimentation rate > 20 mm/hour | 15 (71.4) | 28 (96.6) | 0.04 a |
| Ferritin > 274.66 ng/ml | 9 (42.9) | 23 (79.3) | 0.01 |
| High-sensitivity C-reactive protein >8 mg/liter | 1 (4.8) | 11 (37.9) | 0.01 |
| Interleukin-6> 2.9pg/ml | 18 (85.7) | 28 (96.6) | 0.39 a |
| Interleukin-2> 3.2pg/ml | 1 (4.8) | 8 (27.6) | 0.09 a |
| Interleukin-10> 5.0pg/ml | 2 (9.5) | 6 (20.7) | 0.50 a |
| CD4:CD8 ratio <1.4 | 8 (38.1) | 8 (27.6) | 0.43 |
| SARS-CoV-2 IgM positive | 1 (1 in 12, 8.3%) | 15 (15 in 29, 51.7%) | 0.03 a |
| **Treatment** |  |  |  |
| Corticosteroid | 2 (9.5) | 12 (41.4) | 0.03 |
| Human serum albumin | 5 (23.8) | 18 (62.1) | 0.01 |
| Tocilizumab | 0 | 6 (20.7) | 0.08 a |
| **Chest CT findings** |  |  |  |

|  |  |  |  |  |
| --- | --- | --- | --- | --- |
| Consolidation | 5 (23.8) | 16 (55.2) | 0.03 |  |
| Significant resolution of lung lesion | 11 (52.4) | 6 (20.7) | 0.02 |  |
| Fibrosis | 6 (28.6) | 3 (10.4) | 0.20 | **a** |

**a** represents Chi-Squared Test with continuity correction

Additionally, patients with poor recovery had a higher frequency of corticosteroid therapy and albumin administration than patients with good recovery (Table 2). Odds of poor recovery were higher in patients with older age, hypoproteinemia, hyperlipemia, and corticosteroid therapy (Table 2). As of March 17, 2020, the median time from admission to discharge was 36.5 days (IQR, 24–38 days). A total of 21 (42%) patients had been discharged, whereas 29 (58%) patients were still hospitalized with multifocal lesions on chest CT.

### Virologic characteristics of SARS-CoV-2

A total of 363 viral nucleic-acids tests were performed to explore the viral shedding during the disease course. The viral detection positive rates differed between different specimen types, and varied with day after onset of disease (Supplementary Table 2). The dynamic pattern of viral shedding showed that most patients had intermittent virus shedding. Intriguingly, two patients had significant clinical improvement, but their viral nucleic-acid tests remained positive for a few days. As of March 17, 2020, the median duration of viral shedding was 31 days (IQR, 27–34 days) after symptom onset, and the longest duration of viral shedding was 57 days (Figure 1A). The interval between first and last positive PCR assay results (4 to 22 days) was extensively prolonged in patients with poor recovery.

**Figure 1:**
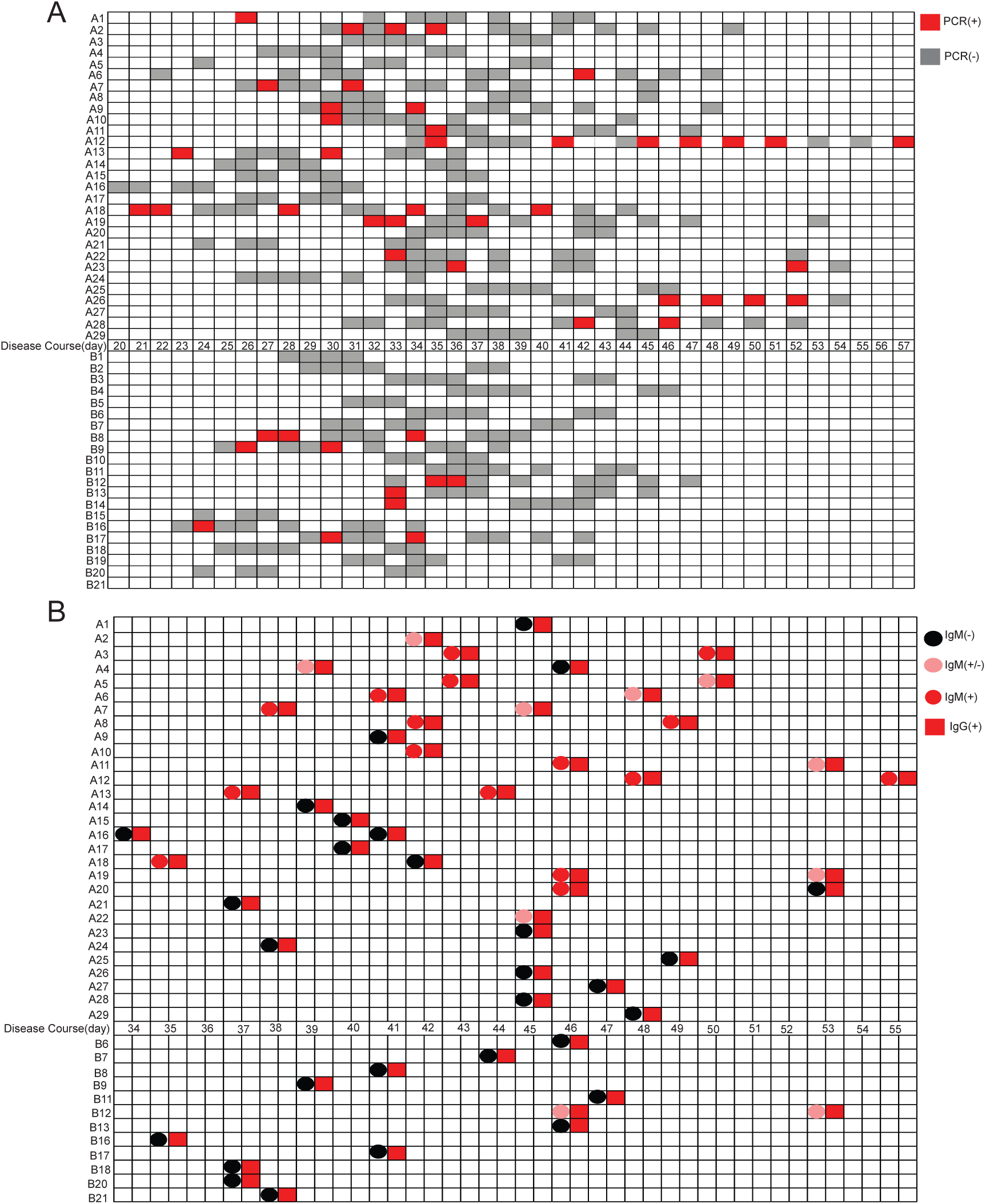
Heat-map shows the virologic characteristics of SARS-CoV-2. In the y axis, A(A1-A29) and B(B1-B21) represents the poor recovery group and good recovery group, respectively. In the x axis, numbers represent the days at symptoms onset. (A) Dynamic viral shedding pattern in the course of disease of patients with severe COVID-19. Red rectangle represents the positive viral nucleic-acid PCR test, while gray rectangle represents the negative viral nucleic-acid PCR test. (B) Serologic tests for SARS-Cov-2 in patients with severe COVID-19 during the disease course. Circle represents IgM test for SARS-CoV-2. In particular, black circle represents negative IgM and pink circle represents weak positive IgM, red represents positive IgM. Red rectangle represents positive IgG test for SARS-CoV-2.

In addition, the serologic tests for SARS-Cov-2 also differed greatly between the two groups. The presence of prolonged SARS-CoV-2 IgM was closely associated with the poor recovery of patients with COVID-19 (p=0.03, Table2). A total of 11 out of 12 (91.66%) patients with good recovery have positive IgG, but negative IgM after hospitalization for one month. In contrast, 15 (51.7%, 15/29) patients with poor recovery had positive test results for both IgM and IgG. As of March 17, 2020, the longest duration of IgM was 55 days at onset of illness, indicating that severe patients with poor recovery were more likely to have a prolonged acute phase of the illness. (Figure 1B).

### Chest CT imaging of severe COVID-19 patients

A total of 195 chest CT imaging results were collected to continuously monitor the disease courses. The most common patterns seen on the initial chest CT scans were bilateral lung involvement (49 out of 50, 98%), ground-glass opacities (GGO) (49 out of 50, 98%), and consolidation (21 out of 50, 42%, Table 1 and Supplementary Figure 1A). Importantly, patients with poor recovery were more likely to have persistent lung damage with diffuse patterns of GGO and multifocal consolidation, and the consolidation pattern was closely associated with the poor outcomes of the patients. (p=0.03, Table 2). The lesions were gradually absorbed with residual GGO, consolidation, interstitial abnormalities, and lung fibrosis observed (Supplementary Figure 3). Most patients in the good recovery group had significant improvement on the chest CT prior to hospital discharge. In contrast, lung lesion resolution was extensively delayed in patients with poor recovery.

Next, several stages were defined to explore the pattern of lung lesion evolution. These included the severest stage, partial resolution, significant resolution, and complete resolution. The most common pattern of evolution was the initial progression to the severest stage, followed by partial resolution and significant resolution. As expected, the duration of the severest stage and lung lesion resolution was markedly extended in patients with impaired lung lesion resolution (Supplementary Figure 1B). In addition, the odds of impaired lung lesion resolution were higher in patients with hypoproteinemia and hyperlipemia. In particular, liver injury, increased levels of TG, and decreased levels of albumin were more commonly seen in patients with impaired lung lesion resolution (Table 3).

**Table 3.** Risk Factors Associated with Lung Lesions Resolution

| **Variable** | **Significant**<br>**resolution**<br>**n = 17**<br>**no. of patients (%)** | **Partial**<br>**resolution**<br>**n = 33**<br>**no. of patients**<br>**(%)** | **P**<br>**value** |  |
| --- | --- | --- | --- | --- |
| **Age** |  |  |  |  |
| ≤ 65 yr | 11 (64.7) | 18 (54.5) | 0.49 | **a** |
| **Coexisting disorder** |  |  |  |  |
| Abnormal liver function | 2 (11.8) | 19 (57.6) | 0.00 |  |
| Hypoproteinemia | 6 (35.3) | 24 (72.7) | 0.01 |  |
| Hyperlipidemia | 8 (47.1) | 25 (75.8) | 0.04 |  |
| **Laboratory findings** |  |  |  |  |
| Leukocyte <4000/mm<sup>3</sup> | 4 (23.5) | 5 (15.2) | 0.73 | **a** |
| D-Dimer > 0.5 mg/liter | 10 (58.8) | 21 (63.6) | 0.74 |  |
| Albumin < 35 g/liter | 6 (35.3) | 24 (72.7) | 0.01 |  |
| Triglyceride > 1.7 mmol/liter | 4 (23.5) | 19 (57.6) | 0.02 |  |
| Lactate dehydrogenase >250 U/liter | 1 (5.9) | 10 (30.3) | 0.10 |  |
| Fibrinogen > 4 g/liter | 8 (47.1) | 15 (45.5) | 0.91 |  |
| C-reactive protein > 8 mg/liter | 4 (23.5) | 17 (51.5) | 0.06 |  |
| Interleukin-4 > 3.2 pg/ml | 3 (17.7) | 17 (51.5) | 0.02 |  |
| Ferritin > 274.66 ng/ml | 7 (41.2) | 25 (75.8) | 0.02 |  |
| hsC-reactive protein > 8 mg/liter | 1 (5.9) | 11 (33.3) | 0.07 |  |
| Interleukin-6 > 2.9pg/ml | 16 (94.1) | 30 (90.9) | 0.88 | **a** |
| Interleukin-2 > 3.2pg/ml | 0 | 9 (27.3) | 0.05 | **a** |
| Interleukin-10 > 5.0pg/ml | 1 (5.9) | 7 (21.2) | 0.32 | **a** |
| CD4:CD8 ratio < 1.4 | 7 (41.2) | 9 (27.3) | 0.32 |  |
| **Treatment** |  |  |  |  |
| Corticosteroid | 4 (23.5) | 10 (30.3) | 0.86 | **a** |
| Intravenous albumin | 3 (17.7) | 20 (60.6) | 0.00 |  |
| Tocilizumab | 0 (0) | 6 (18.2) | 0.16 | **a** |
| **Chest CT findings** |  |  |  |  |
| Bilateral | 16 (94.1) | 33 (100) | 0.34 | **b** |
| Unilateral | 2 (11.8) | 3 (9.1) | 0.84 | **a** |
| Focal | 2 (11.8) | 4 (12.1) | 0.67 | **a** |
| Multifocal | 15 (45.5) | 31 (93.9) | 0.88 | **a** |
| Diffuse | 0 (0) | 5 (15.2) | 0.23 | **a** |
| Peripheral | 9 (52.9) | 13 (39.4) | 0.36 |  |
| GGO | 17 (100) | 32 (97.0) | 1.00 | **b** |
| Centrilobular nodules | 13 (76.5) | 24 (72.7) | 0.96 | **a** |
| Reticular | 0 (0) | 6 (18.2) | 0.16 | **a** |
| Consolidation | 7 (41.2) | 14 (42.4) | 0.93 |  |
| Subpelural line | 4 (23.5) | 8 (24.2) | 0.77 | **a** |
| Pleural effusion | 4 (23.5) | 7 (21.2) | 0.86 | **a** |
| Pleural thickening | 8 (47.1) | 13 (39.4) | 0.60 |  |
| Mediastinal lymphadenopathy | 7 (41.2) | 13 (39.4) | 0.90 |  |
| Fibrosis | 4 (23.5) | 5 (15.2) | 0.73 | **a** |
| Pericardial effusion | 2 (11.8) | 1 (3.0) | 0.55 | **a** |
| **Outcomes** |  |  |  |  |
| Discharge | 11 (64.7) | 10 (30.3) | 0.02 |  |

**a** represents Chi-Squared Test with continuity correction
**b** represents Fisher's Exact Test

### Inflammatory responses in severe COVID-19 patients

To explore the inflammatory-response prolife in severe-condition COVID-19 patients, the laboratory data of inflammatory response-related biomarkers during hospitalization were examined. It was found that most of the patients had ferritin, CRP, ESR, IL-2, IL-4, and IL-6 levels above the normal ranges (Table 1). In contrast, the TNF-α and IFN-γ levels were within the normal range in all of the patients.

Intriguingly, higher levels of CRP, IL-4, ESR, ferritin, and hs-CRP were more often seen in patients with poor recovery (Table 2), and the dynamic inflammatory-response prolife was closely associated with the evolution of lung lesions on the chest CT. Indeed, patients with impaired lung lesion resolution were more likely to have an active inflammatory response indicated by higher levels of ferritin and IL-4 in the serum (Table 3).

Next, to explore the potential role of the inflammatory response in the viral clearance and lung lesion resolution, the clinical details of a patient with the longest duration of viral shedding were examined. In this patient, his viral shedding occurred on day 35, and persistent viral shedding could be observed on days 45–51, indicating active virus replication. Meanwhile, an excess production of cytokine IL-6 was observed on day 36, with rapid progression to the peak stage on day 47. Importantly, extensive lung damage was observed on the chest CT on day 35, but lesions were steadily resolved on days 41–48, with significant improvement on day 56 (Figure 2). A similar pattern could also be observed in other patients (Supplementary Figures 4 and 5). Thus, days of viral shedding and IL-6 concentrations were significantly correlated with the lung lesion evolution on the chest CT in severe COVID-19 patients. It also suggested that patients with poor recovery were more likely to have a prolonged active phase of viral infection. Consequently, proinflammatory responses were promoted, leading to lung lesion resolution and potential viral clearance.

**Figure 2:**
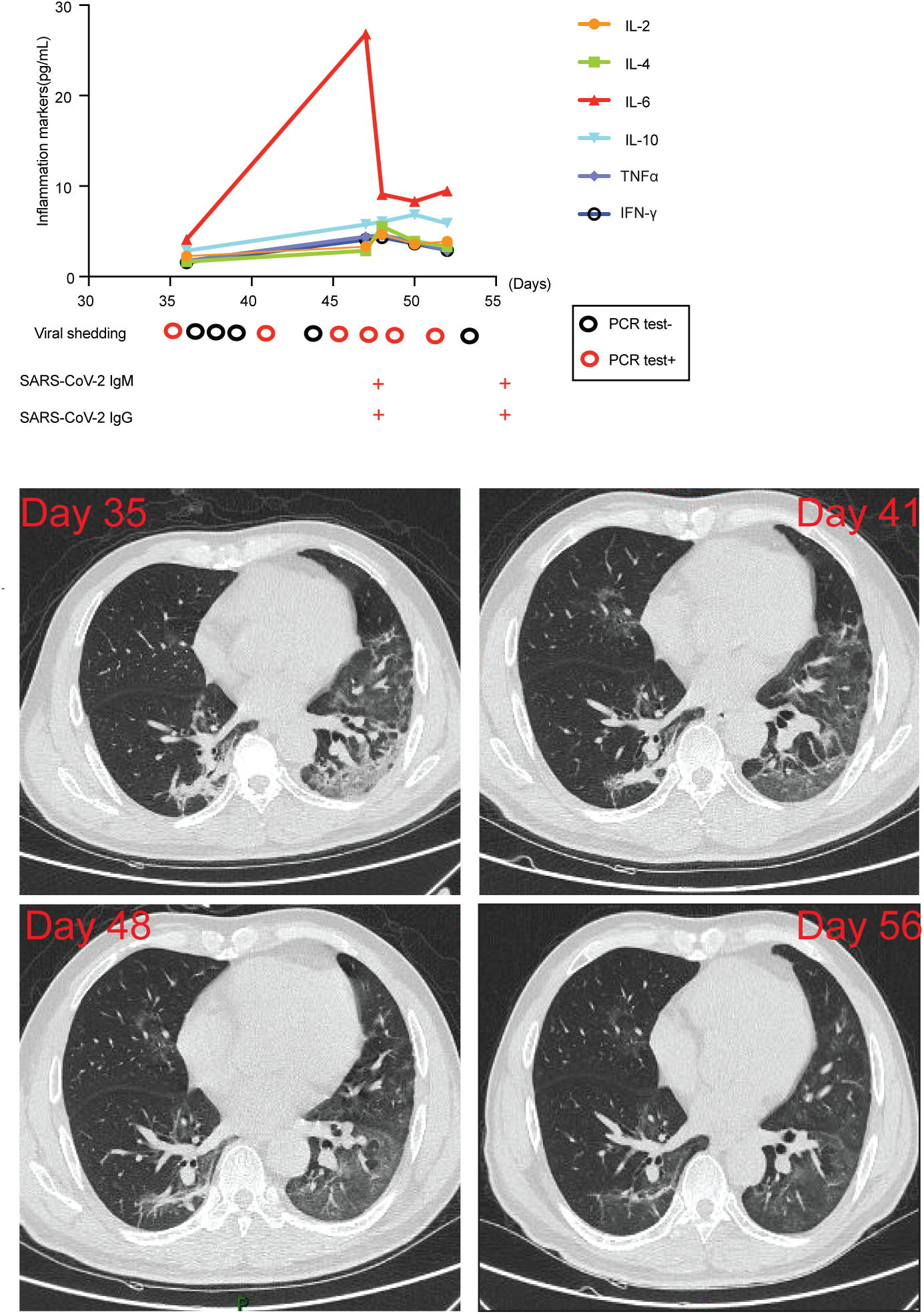
The viral shedding, inflammatory-response and lung lesions resolution prolife in a COVID-19 patient with the longest viral shedding. 55-year-old man, with unknown history of corticosteroid therapy throughout the disease course. Serum cytokine (IL-2, IL-4, IL-6, IL-10, TNF-α and IFN-γ) levels were measured by ELISA assay. Below were the dynamic viral shedding pattern and serological tests for SARS-CoV-2. Black circle represents the negative detection of viral nucleic-acid PCR test, red circle represents the positive result of viral nucleic-acid PCR assay. In the serological tests for SARS-CoV-2, + represents positive for IgM or IgG. Both SARS-CoV-2-IgM and SARS-CoV-2-IgG of serological tests remained positive within day 48-55 after symptoms onset. Serial chest CT images shows the dynamic lung lesions resolution. Day 35 at illness onset: peripheral predominant ground-glass opacity (diffuse pattern) in the left lobe, and partial consolidation in the right lower lobe. Day 41-48 at illness onset: lung lesions steadily resolved. Day 56 at illness onset: significant improvement on chest CT.

## Discussion

Although the lethality of COVID-19 is lower than SARS-CoV (approximately 10%) and MERS-CoV (approximately 34%), COVID-19 rapidly spreads, resulting in a worldwide pandemic^14^. The scope and development of the pandemic are much more severe than SARS and MERS. Compared with the viral shedding at 6–12 days after symptom onset of SARS, the COVID-19 viral shedding is variable and even longer^15^. Moreover, the nucleic-acids of SARS-CoV-2 have been continuously reported to still be detectable in patients who had been discharged after clinical recovery; however, this was not the case for the SARS and MERS nucleic-acids. Therefore, a retrospective, single-center observational study was performed to provide the clinical evidence to determine the virologic characteristics of SARS-CoV-2.

It was found that clinical recovery of patients typically occurred with viral clearing in patients with good recovery. However, it was also found that viral shedding could persist for an interval of few days, despite clinical improvements. Although Zhou et al. reported out that the median duration of viral shedding was 20.0 days (IQR, 17–24 days) in survivors of COVID-19^16^, the duration of viral shedding was found to be greatly extended in this present study, with a median duration of 31 days after symptom onset (IQR, 27–34 days). This difference might be due to specimen collection, disease severity, immunological response, corticosteroid usage, antiviral treatments, and the duration of the observational window in these various populations. A 37-day observational study was conducted, and patient samples were consecutively collected from both the upper respiratory tract and the gastrointestinal tract to improve the detection of SARS-CoV-2 during the course of the disease. This clinical data thus provides convincing evidence of the dynamic viral shedding picture for severely infected patients. In addition, it was found that the interval of viral shedding in patients with poor recovery was markedly prolonged. Importantly, both the IgM and IgG of SARS-CoV-2 in the patients with poor recovery remained positive during the late stage of the disease. This indicated that these patients might have active viral replication in the upper airway, and that persistent viral shedding might be directly linked to the poor recovery of patients with COVID-19. Supporting this notion, it was found that the SARS-CoV-2 RNA could persist until death in non-survivors^17^. Based on the delayed interval of viral shedding pattern and the serologic features of SARS-CoV-2, it is proposed that severe-condition patients with COVID-19 may need at least three negative PCR results with an interval of a few days. In addition, dynamic detection to improve the presence of IgM-negative should also be evaluated to guide discharge planning for COVID-19 patients.

Similar to SARS and MERS, the cytokine profile is closely associated with the severity of COVID-19. Indeed, in this prognostic analysis, it was found that an active inflammatory response was more likely to be triggered in patients with poor recovery, marked by higher concentrations of CRP, ESR, ferritin, and hs-CRP, suggesting that these patients might have a pro-inflammatory factor storm. Previously, Huang et al. reported that the SARS-CoV-2 infection could initiate increased secretion of T-helper-2 (Th2) cytokines (e.g., IL4 and IL10) that suppress inflammation^18^. In this study, it was also found that severe patients with poor recovery had higher levels of IL-4 compared to patients with good recovery. Since the IL-4 levels were slightly elevated in these patients, it was speculated that this might be due to the antagonistic effects of IL-4 during the active phase of the pro-inflammatory response. Importantly, it was found that the wave of the cytokine storm typically occurred during the active phase of viral infection, and a prolonged inflammatory response could be observed in severe patients with poor recovery. Consequently, excessive inflammation damages lung tissue, and subsequent lung repair and regeneration could be initiated. In this regard, lung lesions could gradually resolve and viral clearance might be achieved. Corticosteroids were used frequently for the treatment of severe patients to control the high amounts of cytokines. However, it was found that corticosteroid therapy was closely associated with the poor recovery of severely infected patients. Therefore, corticosteroids should be administrated with great caution if necessary.

Furthermore, viral shedding might persist with the prolonged inflammatory response, leading to persistent lung damage and subsequent pulmonary fibrosis. As of March 17, 2020, 29 patients were still hospitalized with multifocal lesions on the chest CT. A few of them were still in the active phase of viral infection, with intermittent viral shedding and pulmonary fibrosis. It remained unknown whether these patients would develop chronic SARS-CoV-2 infection or be viral carriers. Further follow-up studies in these patients is highly warranted to determine the long-term clinical outcome of severe patients with COVID-19.

In addition to the viral shedding, other risk factors for slower recovery in severe COVID-19 patients were also identified. Consistent with previous studies^19, 20^, it was confirmed that older age, hypoproteinemia, and increased levels of LDH were associated with the poor clinical outcome of COVID-19 patients. In particular, older age, hyperlipemia, hypoproteinemia, corticosteroid therapy, and increased levels of FIB and TG were associated with the poor recovery of COVID-19 patients. Up-to-date information regarding the risk factors associated with lung lesion resolution in severe-condition COVID-19 patients is scarce. The clinical data in this study suggested that all of the patients had lung lesions on chest CT, and the consolidation pattern on chest CT was closely associated with poor recovery. Intriguingly, prognostic factors, including hyperlipemia and hypoproteinemia, were also closely linked to the odds of impaired lung lesion resolution. However, the association of hyperlipidemia and COVID-19 prognosis has not been reported. A previous study reported an association of ACE2 polymorphisms with susceptibility to dyslipidemia^21^. Intriguingly, more than half of the patients in the current study had dyslipidemia. It is speculated that these patients might be particularly prone to SARS-CoV-2 infection, and ACE2 polymorphisms might be linked to patient recovery. Thus, more studies are needed to explore this issue.

This work differs from any other published work, in that we combined the prolife of viral shedding, serological features, inflammatory-response and lung lesions evolutions together, thus providing comprehensive picture of the COVID-19. The prognostic factors of severe patient recovery were identified, which can help clinicians identify patients with poor prognoses at an early stage. In addition, this study provides convincing evidence of the virologic characteristics of SARS-CoV-2, thus it greatly expands our understanding of the natural history of SARS-CoV-2.

Therefore, this study provides helpful management strategies for COVID-19 patients and can help to control the current COVID-19 pandemic.

This study does have some limitations. First, only 50 patients with confirmed COVID-19 were included. It would be better to include a larger number of cases to obtain a more comprehensive understanding of COVID-19. Second, the serologic tests kits for SARS-CoV-2 were available on March 8, 2020, thus the dynamic serologic pattern of SARS-CoV-2 during the early stage was not obtained. Third, the cytokine test results at the early stage of SARS-CoV-2 infection was also not obtained, thus the initial inflammation and immune response profile in severely infected patients remains unclear.

## Data Availability

Availability of all data requests to Dr. Yi Ouyang at the Department of Infectious Diseases, Xiangya Hospital, Central South University, Xiangya Rd 87#. Changsha 410008, P.R.China, or at

## Contributors

YO and JC had the idea for and designed the study and had full access to all data in the study and take responsibility for the integrity of the data and the accuracy of the data analysis. SF, YO, JC, TT, TJ, CZ, SL, HL and YZ contributed to the collection of the clinical data. SF, YO, JC, XF, XS, YH, ZQ, DT and XF contributed to the interpretation of the data. ML, PP, ZX and LX contributed to the analysis and interpretation of chest CT images. SF, YO, JC and XF contributed to writing of the report. YO and JC contributed to critical revision of the report. YS and JM contributed to the statistical analysis.

## Patient and Public Involvement

Patients or the public were not involved in the design, or conduct, or reporting, or dissemination plans of our research.

## Acknowledgments

We thank all our colleagues who helped us during the current study. We greatly appreciate the kind assistance of Liang Peng (Department of Infectious Diseases, 3^rd^ Affiliated Hospital of Sun Yat-sen University, Guangzhou, Guangdong Province, P.R.China) in editing and revising the manuscript, as well as Jie Wei (Department of Health Management Center Xiangya Hospital, Central South University, Changsha, Hunan Province, P.R.China) for her assistance in statistical analysis. We are also grateful to the many front-line medical staff for their dedication in the face of this outbreak, despite the potential threat to their own lives and the lives of their families.

